# Causal attribution fractions, and the attribution of smoking and BMI to the landscape of disease incidence in UK Biobank

**DOI:** 10.1101/2021.12.24.21268368

**Authors:** Anthony J. Webster

## Abstract

Unlike conventional epidemiological studies that use observational data to estimate “associations” between risk factors and disease, the science of causal inference has identified situations where causal estimates can be made from observational data, using results such as the “backdoor criteria”. These results are combined here with established epidemiological methods, to calculate simple population attribution fractions that estimate the causal influence of risk factors on disease incidence, and can be estimated using conventional proportional hazards methods. A counterfactual argument gives an attribution fraction for individuals. Causally meaningful attribution fractions cannot be constructed for all risk factors or confounders, but they can for the important established risk factors of smoking and body mass index (BMI). Using the new results, the causal attribution of smoking and BMI to the incidence of 226 diseases in the UK Biobank are estimated, and summarised in terms of disease chapters from the International Classification of Diseases (ICD-10). The diseases most strongly attributed to smoking and BMI are identified, finding 11 with attribution fractions greater than 0.5, and a small number with protective associations. The results provide new tools to quantify the causal influence of risk factors such as smoking and BMI on disease, and survey the causal influence of smoking and BMI on the landscape of disease incidence in the UK Biobank population.

## Introduction

This article has two aims: (i) To derive simple formulae for estimating causal influences of risks such as smoking using observational data and conventional proportional hazard estimates. (ii) To estimate the causal influence of smoking and BMI on patterns of disease incidence. Conventional epidemiological methods are unable to use observational data to make causal claims, and graphical arguments from causal inference theory [1] suggest that it may not always be possible to make causal estimates of all quantities using observational data. However, it is shown here that for important risk factors such as smoking and BMI, there are reasonable assumptions about causal processes (figure 1), that allow results from causal inference [1] to be used to calculate causal estimates from observational data. Furthermore, it is shown how these can be evaluated using conventional epidemiological methods, making the results accessible to epidemiologists who are unfamiliar with results from causal inference. The new formulae are easy to understand and evaluate, and are used here to estimate how smoking and BMI modify the patterns of disease incidence in the UK Biobank dataset. To the author’s knowledge, this is the first attempt to use methods from causal inference to estimate the causal influence of established risk factors on the overall incidence of diseases in a population.

**Figure 1:**
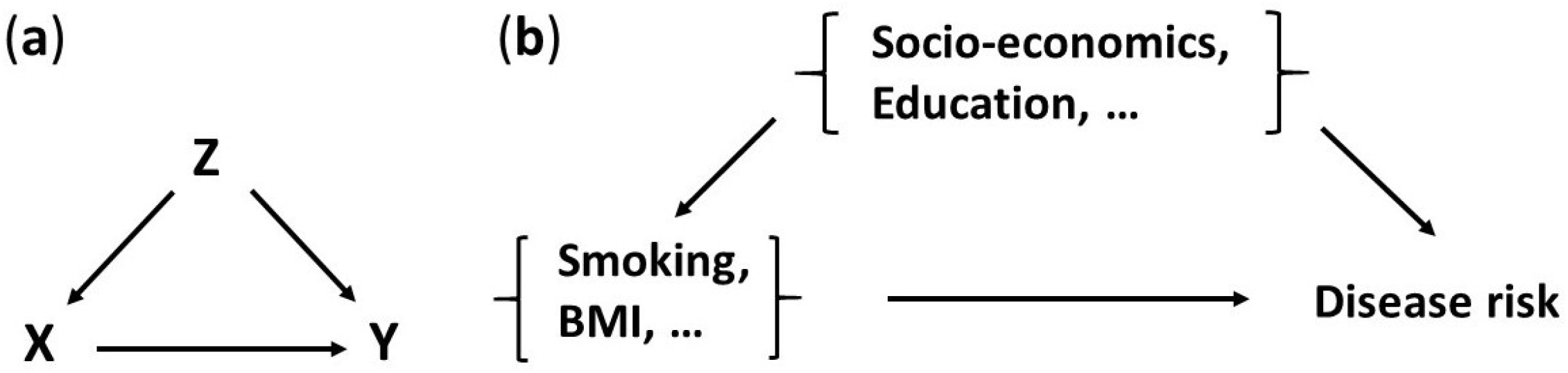
Consider the influence of one or more exposures *X*, on diseases *Y*, with confounding variables *Z* that satisfy the “backdoor criteria” [1, 7] (figure a). For example, *X* might include BMI, alcohol, and smoking, with confounders *Z* of socio-economic status and education (figure b).

The original aim of this work was to quantify how risk factors modify the overall patterns of disease that are observed in UK Biobank [11], and to use this information to improve the classification of diseases. Unfortunately, conventional epidemiological methods are unable to use observational data to make causal claims, and instead describe results in terms of “associations”. Associations between potential risk factors and diseases are usually quantified with attributable fractions and relative risks [2, 3], that are estimated using proportional hazard models [3, 4]. However, there are several ways of defining and estimating attributable fractions [3, 5, 6], and relative risks do not generally have a causal interpretation [3].

The science of causal inference [1, 7–9] has recently identified circumstances where causal estimates are possible using observational data. These include results such as the “backdoor criteria” and methods involving the “do” calculus [1, 7], that are combined here (in the Methods), with estimates of relative risks from conventional epidemiological studies [4], to allow causal estimates using observational data. For a short introduction to causal inference see Pearl et al. [7], that summarises several of the key results from the comprehensive text by Judea Pearl [1]. The Supplementary Material contains additional results to those in the Methods, that consider unmeasured confounders using the “frontdoor” criteria from causal inference [1, 7], and use proportional hazard estimates to relate the results to those from mediation analyses [9]. In the Methods a population attribution fraction is derived, that estimates the proportional change in disease incidence caused by a risk factor, and shows how it can be estimated using conventional proportional hazard studies. The attribution fraction can be used when estimates are of causal associations, in the sense outlined in the Methods and illustrated in figure 1. It is closely related to the average causal effect (ACE) [1, 7], and can (in principle), agree with conventional attribution fractions when these are combined with estimates of causal associations [2, 5]. In the Discussion, an attribution fraction for an individual is formulated using a counterfactual argument for the “effect of treatment on the treated” [1, 7], that gives a simple and well-known expression in terms of an individual’s relative risk. Unless stated otherwise, the rest of this article will use “attribution fraction”, to refer to the population attribution fraction.

The following Methods section derives the attribution fractions that are used, and describes the survival analyses that are used to estimate them. Only established risk factors and confounders were considered, with a causal diagram assumed as in figure 1. It seemed reasonable to assume (figure 1), that confounders such as socio-economic status influence both disease risk and the presence of risk factors such as smoking status or BMI, but the main influence of risk factors such as smoking or BMI are directly on health. The Results section summarises the overall results in terms of attribution fractions and International Classification of Disease (ICD-10) chapters, and identifies diseases with the largest (and smallest), attribution fractions. The process of selecting diseases for study is detailed elsewhere [10], along with further information on the UK Biobank data that was used [10, 11]. Using the newly derived attribution fractions, a conventional epidemiological study was used to estimate the proportion of disease that should be attributed to smoking or BMI, for 226 diseases in UK Biobank. The results emphasised the heterogeneous influence of risk factors, that ranged from protective associations for several diseases, to 11 diseases whose attributable fractions exceeded 0.5. Diseases were characterised by their attribution fractions, that allowed them to be ranked and classified in terms of their risk modifiability in terms of smoking and BMI. Although it is possible that estimates can be improved by individualised studies of each disease, this study accounts for well-known established factors, while allowing a broad survey of the overall influence of smoking and BMI on disease. The Discussion mentions some important related results, discusses the consequences of the Results, and summarises the limitations of the study.

## Methods

### Modelling interventions with the “backdoor criteria”

Figure 1 shows the causal relationships between potential risk factors *X* and confounding factors *Z*, that are assumed here. The confounding factors *Z* are assumed to include education and socio-economic status, and the risk factors *X* included smoking, BMI, height, and alcohol consumption, and for women only, HRT use and parity. The presence or absence of disease is indicated by *Y* = 1 or *Y* = 0. For this causal model (figure 1), it is possible to estimate the consequences of setting BMI, alcohol, and smoking to a specific value *X* = *x*, corresponding to do(*X* = *x*) using the “do” notation of Pearl [1, 7]. The situation is described by the well-known “adjustment” formula [1, 7], that states,

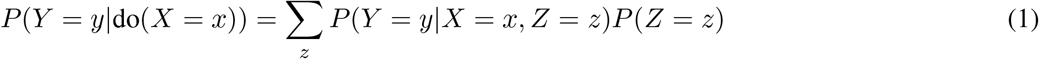

where for continuous variables the sums are treated as integrals, upper case *X, Y, Z* correspond to specific values of random variables, and lower case *x, y, z* can take any allowed value. The formula accounts for the confounding influence of *Z* on both *X* and disease risk, and differs from the equivalent result from conventional probability theory [12], that would have *P* (*Z* = *z*|*X* = *x*) instead of *P* (*Z* = *z*). With *Y* = 1 denoting the presence of disease, then,

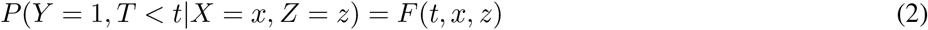

where *F* (*t, x, z*) is the distribution function for disease onset within time *t*, and the covariates are (a vector) of risk factors *x* and (a vector) of confounding factors *z*. For a probability density *f* = *dF/dt* and hazard function *h* = *f/S*, then the cumulative hazard function is 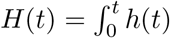, and a proportional hazards model will assume that 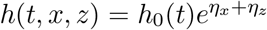, where *h*_0_ is the baseline hazard function with linear predictors *η*_*x*_, *η*_*z*_ for the risk factors and confounders respectively [4] (the linear predictor function is sometimes referred to as the “linear compoent”, “risk score”, or “prognostic index”). With these assumptions,

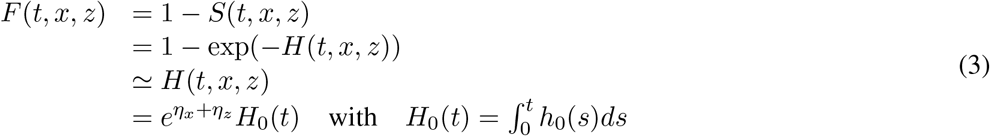

where in going from the 2nd to 3rd line we assume sufficiently rare diseases that exp(− *H*(*t, x, z*)) ≃ 1 − *H*(*t, x, z*), as is the case for the first diagnosis of most diseases in UK Biobank [11, 13] (more generally *F* (*t, x, z*) ≤ *H*(*t, x, z*)), and in going from the 3rd to the 4th lines we assume that the proportional hazards assumption [4] is valid for the disease being studied. The approximations are explored further in the Discussion and Supplementary Material. Now using Eq. 1,

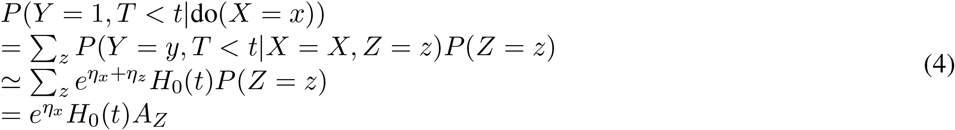

with 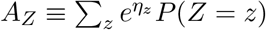, and sums replaced by integrals for continuous variables. This allows the incidence rates to be calculated for a (possibly hypothetical) situation where we have intervened in some way to set *X* = *x*, in terms of a baseline hazard function that is estimated in the usual way using observational data, in which *Z* can be correlated with both *X* and disease risk. Note that *P* (*Z* = *z*) and *P* (*X* = *x*) are implicitly the population values at the study’s start. At the baseline values of *x* = *x*_0_ and *z* = *z*_0_, by definition *η*_*x*_(*x*_0_) = 0 and *η*_*z*_(*z*_0_) = 0, so Eq. 4 gives *P* (*Y* = 1, *T < t|X* = *x*_0_, *Z* = *z*_0_) = *H*_0_(*t*).

### Attributable fractions

Attributable fractions are intended to describe the proportion of disease incidence that is caused by an exposure, or can be avoided by an intervention. They can be defined in several related but distinct ways [3, 6]. Here the attributable fraction for the situation described by figure 1 is considered. To consider the causal influence of a subset *X*, of risk factors *X* and *W*, the risk factors are considered to be composed of both *X* and *W*. If *P* (*Y* = 1, *T < t*) is the probability of observing a disease at age *T*, less than *t*, then the average causal effect of risk factors on disease risk in a population compared with baseline risk factors is, *P* (*Y* = 1, *T < t*) *− P* (*Y* = 1, *T < t*|do(*X* = *x*_0_)), and the excess fraction is,

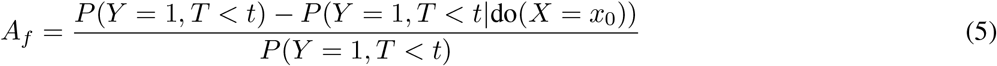

In this definition *A*_*f*_ is in principle a function of age *t*, but in the examples here the dependence on age is usually small, and an age-independent definition will be suggested later. The numerator of 5 is the average causal effect (ACE) [1] of the risk factors *X* on the population’s disease risk, compared with the baseline values *X* = *x*_0_. It is divided by the probability of risk in the population, giving an excess risk fraction, that is referred to here as an attributable fraction. To evaluate this, firstly note that,

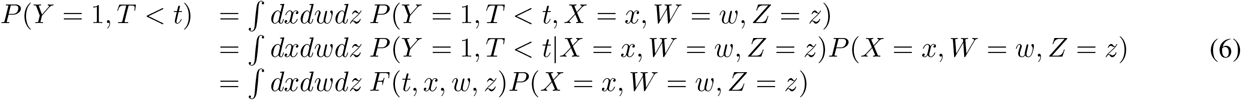

where integrals should be replaced by sums for non-continuous variables. *P* (*Y* = 1, *T < t* | do(*X* = *x*_0_)) can be evaluated similarly, and for situations described by figure 1, the backdoor adjustment formula is used in the second line below,

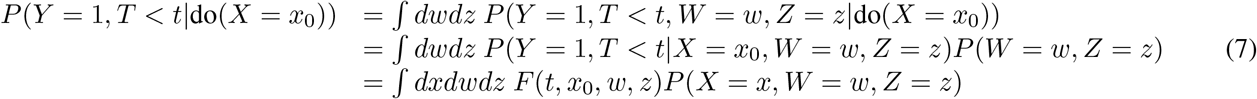

With the only difference from Eq. 6 being the replacement of *x* with *x*_0_. Therefore, using Eqs. 6 and 7, the excess fraction is,

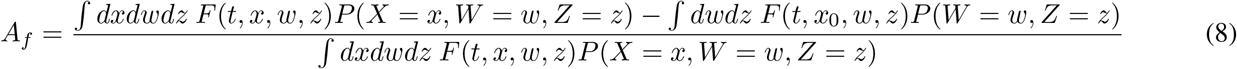

Eq. 8 for *A*_*f*_ becomes independent of age *t* as *t* → 0. This offers an age-independent definition of *A*_*f*_ that is an unobservable theoretical limit, but can be evaluated using estimated survival curves such as Weibull distributions [13] or the approximations outlined below. In practice, for the epidemiological study here using a proportional hazards model, for most diseases in most individuals, the estimated incidence rates were sufficiently low that *A*_*f*_ ≃ *A*_*f*_ (*t* = 0) for most of a typical UK human lifespan.

The integrals in 8 can be estimated by noting that 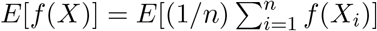 and that the variance 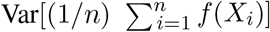 (1/*n*)Var(*f*(*X*)) → 0 as *n* → *∞*. This allows the integrals to be approximated by a sum over the observed data, which is reasonable if the number of data points is sufficiently large in each level of categorical data considered. For example, in the study of UK Biobank described later with nearly 500,000 participants, the smallest category was for current smokers, but this included over 50,000 smokers. With this approximation,

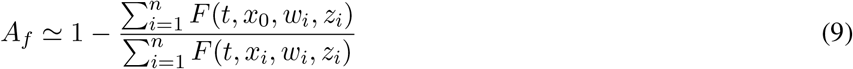

that can be evaluated using numerical fits for the incidence of disease, such as those using a Weibull distribution [13], but will be a function of age *t*. Eq. 9 can be simplified further by assuming that the data can be described by a proportional hazards model, and approximating [13] 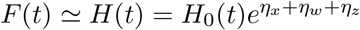, where *η*_*x*_, *η*_*w*_, *η*_*z*_ are linear predictors respectively involving *x, w*, and *z*. This gives,

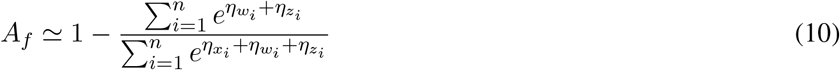

which is independent of age *t*, and easily evaluated using conventional proportional hazards modelling. Eq. 10 might alternately be written as,

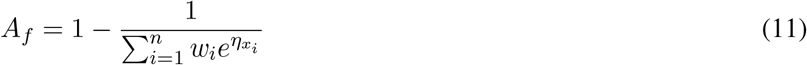

with,

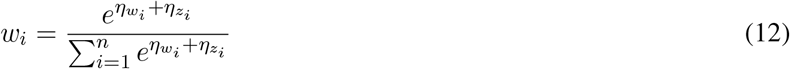

which shows that the relative risk is weighted by the influence of confounders and other risk factors, but is otherwise similar to conventional attributable fractions that have 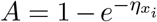. When there are no confounders *z* or other risk factors *w*, then the terms in Eq. 12 become 1, and *w*_*i*_ = 1*/n*, so that 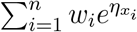 is just the average of 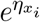 across the population. Eq. 11 shows that if the relative risk 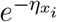 is positively correlated with the relative risks from confounding and other potential risk factors 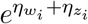, then 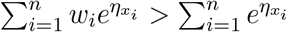. and the attribution fraction will have been increased by accounting for the confounding and other potential risk factors.

For comparison, the World Health Organisation (WHO) uses an attributable fraction that is defined as [2],

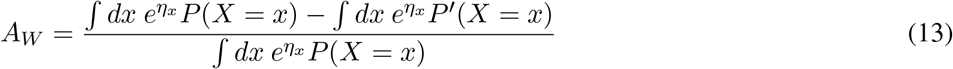

where *P*^*′*^(*X* = *x*) is an alternative probability distribution for *X*. If we take *P*^*′*^(*X* = *x*) to be a delta function centred on *X* = *x*_0_, with 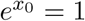, so that we are comparing the population with one where *X* = *x*_0_, then,

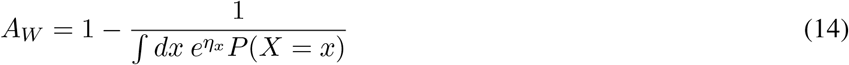

which is the same as would be obtained by assuming that 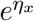 and 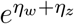 are uncorrelated in Eq. 10. The Supplementary Material (A.1), shows that the *A*_*W*_ provides a lower (upper) bound on *A*_*f*_ if 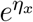 is positively (negatively) correlated with 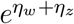. In general Eqs. 8 and 13 will differ, and neither should have a causal interpretation unless the causal model satisfies suitable conditions such as those in figure 1 that ensure that causal associations are being estimated.

To compare the attributable risk between setting *X* = *x*_1_ and *X* = *x*_2_, the equivalent expression to Eq. 10 is,

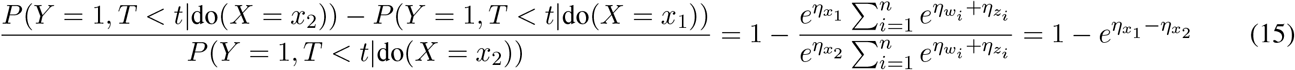

which is just the conventional result for attributable fraction in terms of the relative risk.

The analysis here will apply more generally than to studies involving disease, or health. Similar results will apply whenever *F* (*t, x, z*) can be factored as *H*_0_(*t*)*g*(*x*)*q*(*z*), for some functions *g*(*x*) and *q*(*z*), as was possible here because we consider a proportional hazards model and situations where the incidence is sufficiently rare that we can approximate *F* (*t, x, z*) ≃ *H*(*t, x, z*).

### Number of attributed cases

The proportion of disease cases that are attributed to a risk factor is only important if the disease is sufficiently common. The change in the number of cases of disease can be estimated using the estimated attributable fraction and the number of observed cases of disease. If *N* is the population size under consideration, and we define *P ≡ P* (*Y* = 1, *T < t*), and *P*_0_ *≡ P* (*Y* = 1, *T < t*|do(*X* = *x*_0_)), then,

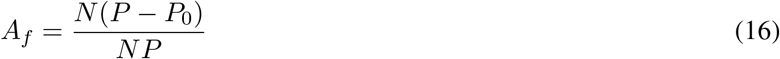

If we approximate *NP* as the observed number of cases in the population being studied *N*_*obs*_, then we can estimate the number of extra (or fewer) cases from the attributable fraction *A*_*f*_, with,

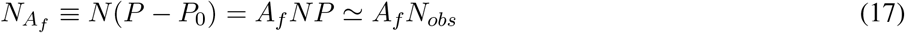

This gives a simple estimate for the number of cases that are attributable to a risk factor. However, this is the number of attributable cases of hospital admissions, for diseases included by the study’s selection criterion - first admissions in an ICD-10 chapter in this paper. This latter estimate could substantially differ from our perception of the number of hospital admissions caused by a specific disease, that could be dominated by sequences of hospital visits, or result from a different original underlying cause. For that reason, attributable fractions are generally a better measure of the causal influence of risk factors on the risk of disease.

If the attributable fraction given by Eq. 10 were negative, then instead of considering (*P* − *P*_0_)*/P*, an alternative would be to consider (*P*_0_ − *P*)*/P*_0_. However, provided *A*_*f*_ is reasonably small, then the two estimates have approximately the same magnitude, with a change in sign to indicate the direction of effect. Expanding (*P*_0_ − *P*)*/P*_0_ in terms of *A*_*f*_ = (*P* − *P*_0_)*/P*, gives,

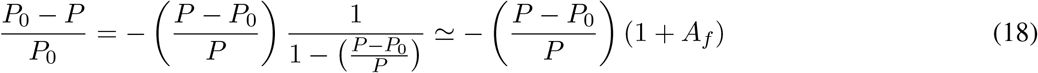

Showing that both expressions are approximately equal in magnitude if *A*_*f*_ is small.

### Survival analysis

To minimise the potential for confounding by prior disease, only the first incidence of disease in each ICD-10 chapter was considered for each individual. Diagnoses that were the primary cause of hospital admission were considered. These will have passed a threshold of severity to trigger hospital admission, and are recorded with an ICD-10 code in hospital episode statistics (HES). Individuals who reported diabetes at entry to the study were excluded, to ensure that any new cases of diabetes would almost entirely involve type II diabetes. For each disease, the participant’s data were excluded if onset occurred before they entered the study, or if they had a prior hospital diagnosis of cancer other than non-melanoma skin cancer. The incidence rates of the diseases considered are “rare” in the approximate sense used to estimate attribution fractions [13]. A survival analysis using age as the time variable was left-truncated at a participant’s entry to the study, right-censored if there was: death, cancer other than non-melanoma skin cancer, or the study period ended. All diagnoses recorded between entering the study and 31st January 2020 were included, as recorded in UK Biobank HES data on 8th December 2021. Data beyond 31st January 2020 were likely to be influenced by the COVID-19 pandemic and were omitted. Analyses were multiply adjusted using a proportional hazards model, with men and women studied separately, and a causal model assumed as in figure 1. Adjustment considered the established risk factors of: smoking status (never, previous, or current), alcohol consumption (rarely - less than 3 times per month, sometimes - less than 3 times a week but more than 3 per month, regularly - 3 or more times each week), education (degree level, post-16 but below degree, to age 16 or unspecified), socio-economic status (tertiles), height (sex-specific tertiles), body mass index (BMI) (sex-specific tertiles), and for women we also adjusted for: hormone replacement therapy (HRT) use ever (yes, no), and one or more children (yes,no). Baseline was taken as: never smoker, rarely drink, brisk walking pace, degree-level education, minimum deprivation tertile, minimum height tertile in men (or women), middle BMI tertile in men (or women), and women with no children or HRT use. Only diseases with at least 140 cases were considered. This ensured there were at least 10 cases per parameter to adjust from baseline, even if a parametric e.g. Weibull model with an extra two parameters to fit the baseline hazard function were considered [13]. Sensitivity analyses excluded participants with a broader range of prior diseases, leading to fewer total cases and fewer diseases included in the study. There were less than 1% missing values, allowing a complete case analysis. Numerical work and plots used R [14], and packages used here included: survival[15] and grr[16].

Attribution fractions for the UK Biobank population were considered for three situations: observed population verus never-smoked, observed population versus middle BMI tertile, and observed population versus never-smoked and middle BMI tertile. The latter case is comparing the correlated exposures of BMI and smoking status in the observed population, to a situation where BMI and smoking are set to their baseline values of never-smoked and middle BMI tertile. Because the baseline BMI tertile is the middle tertile, current smokers could be correlated with either the top or bottom BMI tertile. Frequency of alcohol consumption was adjusted for but not studied, because it is a less precise measure than smoking status or BMI, and it is known to have inconsistent associations with disease risk in different studies [17].

## Results

UK Biobank data [11] was used to estimate the attribution of smoking and BMI to the incidence of over 400 hospital diagnosed diseases in men and women. Diseases were characterised by their attribution fractions, allowing them to be ranked and classified in terms of the modifiability of population risk, by a change of smoking status or BMI. Information on the selection of diseases for study is detailed elsewhere [10], as are details of the UK Biobank cohort [10, 11]. Although the survival analyses could be improved by individual study of each disease, the study here accounts for the strongest established risk and confounding factors, while allowing a broad survey of the overall influence of smoking and BMI on disease in the UK Biobank cohort.

Plots and tables include diseases with statistically significant associations with current smoking versus never smoked, or maximum versus middle BMI tertile, after an FDR multiple-testing adjustment. Where results involve both smoking and BMI then diseases were included if they are included in either of the smoking-only, or BMI-only results. This left 129 diseases associated with BMI, 153 diseases associated with smoking, and 226 diseases that were associated with either smoking or BMI. To explore the sensitivity of the estimates to the strength of confounding factors, estimates made using Eqs. 10 and 13 were compared (supplementary figure 2). As expected, the influences of confounding are more noticeable for smaller attributable fractions, but even in those cases, the estimates rarely differ by more than about 20%. With a handful of exceptions, such as Parkinson’s disease (G20), estimates with Eq. 10 were larger than with 13, as would be expected if the influence of smoking and BMI were positively correlated with the influence of the confounding factors in the model.

Figure 2 shows the median attributable fractions for the combined influence of smoking and BMI on the incidence of disease in each ICD-10 chapter, with the width of the bar plots proportional to the number of diseases in the estimate. Diseases of the respiratory system (X) have the largest median attribution fraction, of about 0.3, closely followed by endocrine, nutritional, and metabolic diseases, that are both almost double the next largest values. Diseases of the skin and subcutaneous tissues (XII) and of the nervous system (VI), both have median attribution fractions near 0.15. Neoplasms and circulatory diseases account for 22% of all the diseases, and have the next largest median attributable fractions. There are seven chapters with median attribution fractions greater than 0.1, and these include 100 diseases, 50 of which are neoplasms and diseases of the circulatory system.

**Figure 2:**
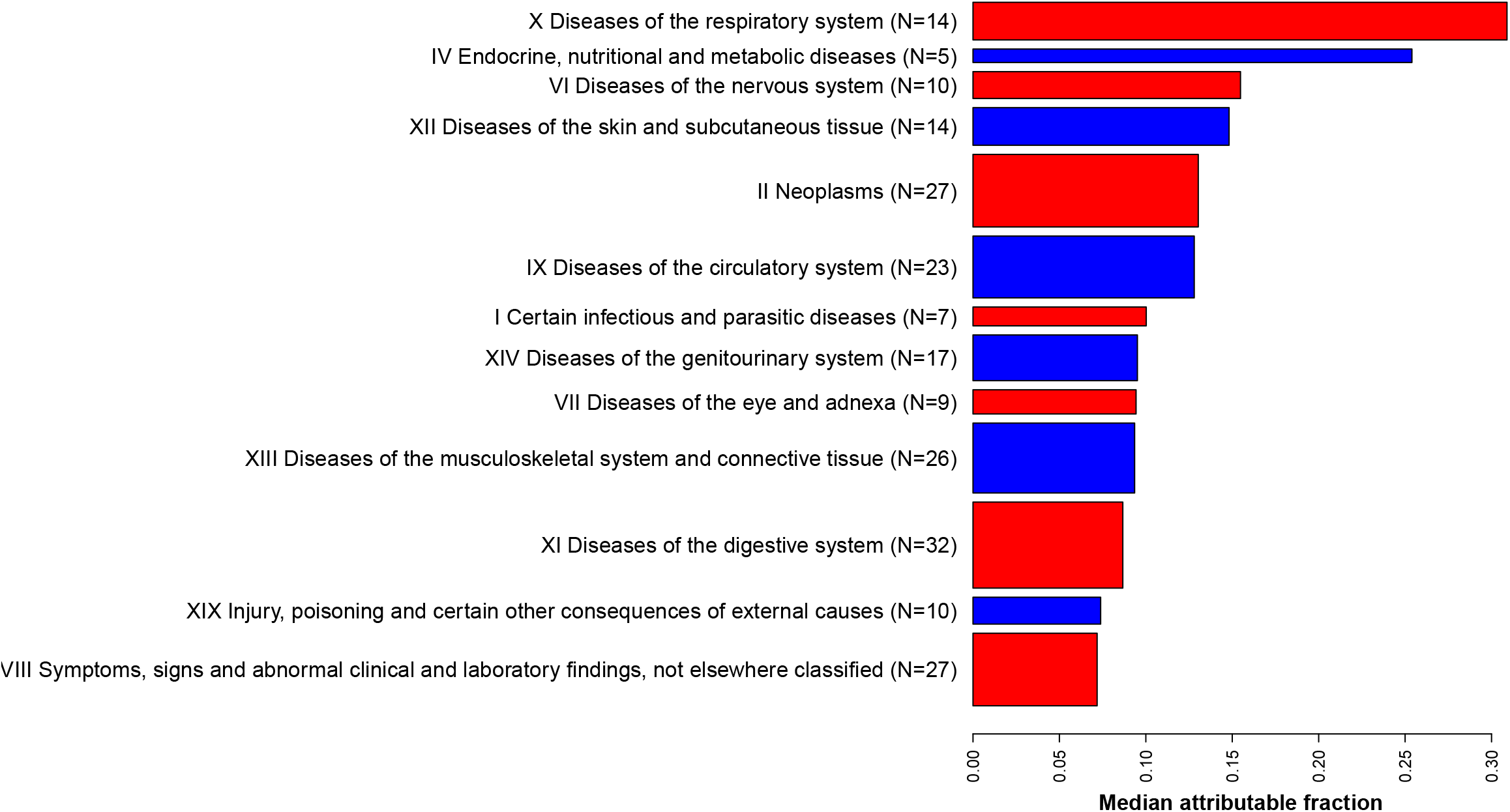
Median attributable fractions for each ICD-10 chapter with at least 5 diseases where *A*_*f*_ *>* 0.2. Bar widths are proportional to the number of diseases in each chapter.

The 26% of diseases that had *A*_*f*_ *≥* 0.2 are listed in table 1. There are 11 diseases with *A*_*f*_ *≥* 0.5 and 21 with *A*_*f*_ *≥* 0.35. Given the limitations of the analysis and the potential for regression dilution bias, it is possible that more than 11 diseases could have *A*_*f*_ *≥* 0.5. For diseases with more than half the cases attributed to smoking and BMI, it seems reasonable to regard the influence of smoking and BMI on the population as “pathogenic”, in a similar way that strong genetic risk factors are often described as pathogenic. One third of the 226 diseases had an attributable fraction with | *A*_*f*_ | *>* 0.17, and two thirds had | *A*_*f*_ | *>* 0.06. Although the mean attribution fraction for the combined influence of smoking and BMI was 15%, the estimated attributable number of extra cases was only ≃ 8%, reflecting the fact that the most common diseases (with the most cases), tended to have smaller attributable fractions.

**Table 1:**
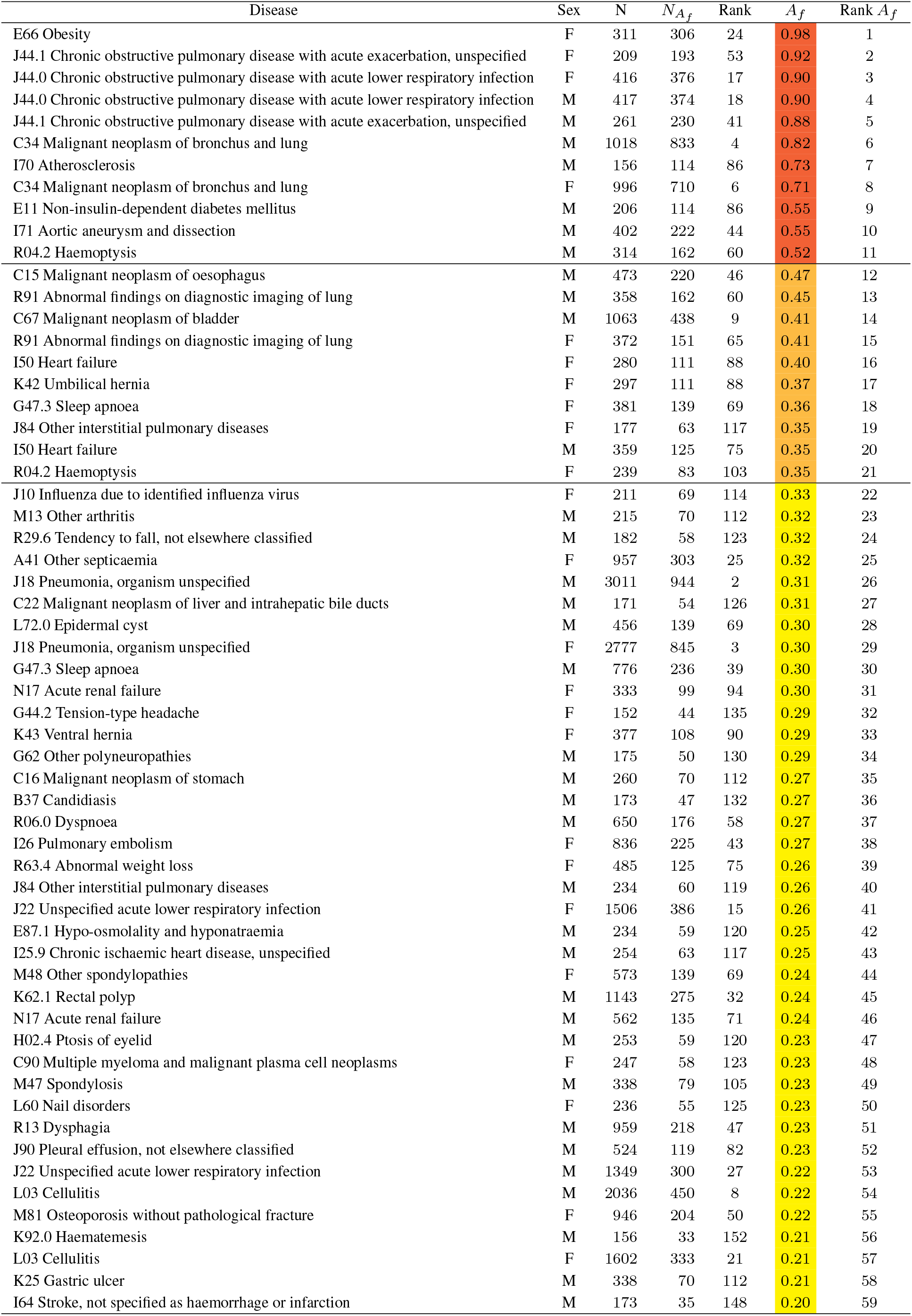
Attributable fractions *A*_*f*_ for both smoking and BMI are estimated with Eq. 10, ranked, and listed if *A*_*f*_ *≥* 0.2. Colours: *A*_*f*_ *≥* 0.5 (red), 0.5 *> A*_*f*_ *≥* 0.35 (orange), 0.35 *> A*_*f*_ *≥* 0.2 (yellow). Sex: diseases in males (M) or females (F), *N* : total cases, 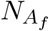: attributed cases.

Diseases were ranked in terms of their attribution fractions for smoking and BMI (figure 3). Figure 3 identifies an important point, that even established risk factors such as smoking and BMI can have protective associations with some diseases. The 20 diseases that smoking and BMI have the strongest protective associations with are listed in table 2. There were 12 diseases whose protective association had an attributable fraction with magnitude greater than 0.1, and 3 with magnitude greater than 0.2. Melanoma in situ (D03), had the strongest protective association of -0.29, where the sign is taken to indicate the direction of effect as discussed in “Number of attributed cases”.

**Figure 3:**
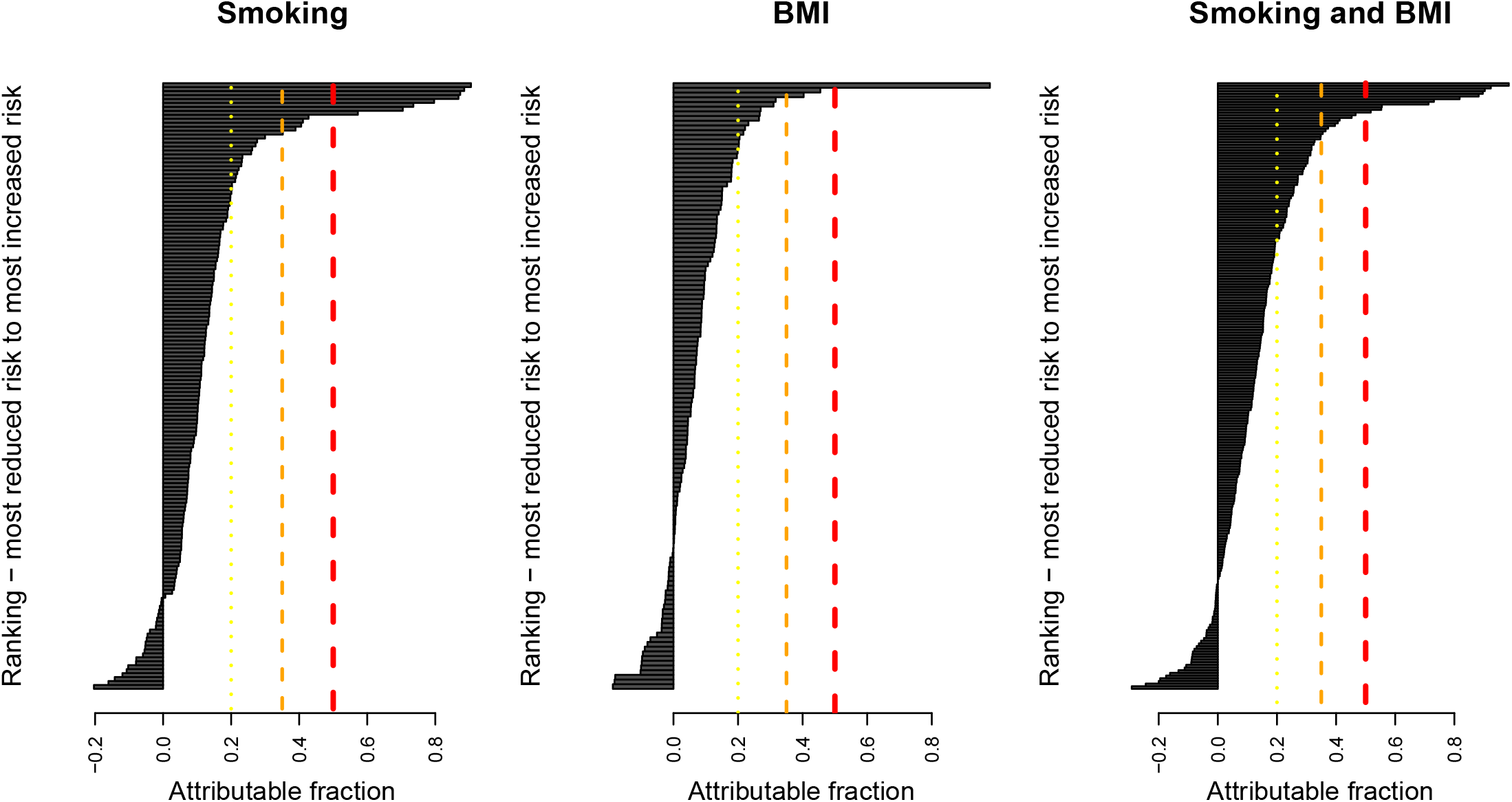
For diseases with a statistically significant association with smoking or BMI after an FDR multiple-testing adjustment, attributable fractions *A*_*f*_ were calculated with Eq. 10. Lines indicate *A*_*f*_ = 0.5 (red), *A*_*f*_ = 0.35 (orange), *A*_*f*_ = 0.2 (yellow). *A*_*f*_ *<* 0 indicates a protective association.

**Table 2:** Diseases with the strongest protective associations, ranked by the proportion of disease attributed to a combination of smoking and BMI (*A*_*f*_). Sex indicates diseases in males (M) or females (F), *N* are total cases, 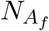 are the number of cases attributed to smoking and BMI, *A*_*f*_ is the attributable fraction.

| Disease | Sex | N | $N_{A_f}$ | Rank | $A_f$ | Rank $A_f$ |
| --- | --- | --- | --- | --- | --- | --- |
| D03 Melanoma in situ | M | 272 | -79 | 14 | -0.290 | 1 |
| K31.7 Polyp of stomach and duodenum | F | 870 | -212 | 9 | -0.240 | 2 |
| N41 Inflammatory diseases of prostate | M | 572 | -114 | 10 | -0.200 | 3 |
| N81 Female genital prolapse | F | 4199 | -819 | 1 | -0.190 | 4 |
| R79 Other abnormal findings of blood chemistry | M | 1905 | -333 | 5 | -0.170 | 5 |
| S02 Fracture of skull and facial bones | M | 355 | -57 | 18 | -0.160 | 6 |
| C43 Malignant melanoma of skin | M | 723 | -96 | 11 | -0.130 | 7 |
| S76.1 Injury of quadriceps muscle and tendon | M | 215 | -24 | 25 | -0.110 | 8 |
| M20.1 Hallux valgus (acquired) | F | 2875 | -307 | 6 | -0.110 | 9 |
| C61 Malignant neoplasm of prostate | M | 5800 | -521 | 2 | -0.090 | 10 |
| B34 Viral infection of unspecified site | M | 319 | -28 | 23 | -0.089 | 11 |
| N40 Hyperplasia of prostate | M | 3928 | -344 | 4 | -0.088 | 12 |
| M16 Coxarthrosis [arthrosis of hip] | M | 3167 | -272 | 7 | -0.086 | 13 |
| J90 Pleural effusion, not elsewhere classified | F | 327 | -28 | 23 | -0.084 | 14 |
| K40 Inguinal hernia | F | 493 | -39 | 20 | -0.079 | 15 |
| R19.8 Other specified symptoms and signs involving the digestive system and abdomen | F | 302 | -22 | 27 | -0.072 | 16 |
| C44 Other malignant neoplasms of skin | M | 6095 | -391 | 3 | -0.064 | 17 |
| K31.7 Polyp of stomach and duodenum | M | 300 | -17 | 31 | -0.057 | 18 |

### Sensitivity analysis

Participants with hospital reports of prior cancers other than non-melanoma skin cancers were excluded from the main study, but self-reported cancers or other prior diseases were not. It is possible for example, that a heart attack might be followed by weight loss, and including participants with prior heart attacks could weaken a potential association between BMI and heart disease. In contrast, smoking might increase the risk of some diseases for which a substantial proportion occur before entry into the UK Biobank study. In that case, including participants with the prior disease might strengthen the associations. The question of how best to study sequences of disease is an example where causal understanding is not enough, and new statistical methods or data are likely to be required. It might be an intractable question, due to the vast possible combinations of sequences of 100s of diseases, and it is further complicated by the complex time-dependent exposures and accumulation of genetic mutations that any individual experiences. Therefore a senstivity analysis compared the paper’s main results with a second analysis that excluded participants who had reported any cancer other than non-melanoma skin cancer, or any serious cardiovascular disease of heart disease, stroke, arterial or pulmonary embolisms, or subarachnoid haemorrhage.

Differences between the main study and the sensitivity analysis were small. There were six diseases whose attribution fractions increased from *A*_*f*_ < 0.2, to *A*_*f*_ ≥ 0.2, these are listed in table 3. The difference between attribution fractions in the two studies had a mean and median of -0.006 and -0.005 respectively, and a standard deviation of 0.029. The differences in magnitude were typically equivalent to about 10%. The attribution fractions of six diseases changed by more than 0.05. These included increased attributable fractions for: I50 - heart failure in women (0.40 to 0.48), R29.6 - tendency to fall in men (0.32 to 0.41), and decreases in: C16 - stomach cancer in men (0.27 to 0.21), J10 - influenza in women (0.33 to 0.27), J22 - lower respiratory infections in men (0.24 to 0.17), and N17 - acute renal failure (0.24 to 0.17).

**Table 3:** The sensitivity analyses found six additional diseases with *A*_*f*_ *≥* 0.2, for the combination of both smoking and BMI, that would have appeared in table 1. Sex: diseases in males (M) or females (F), *N* : total cases, 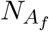 : attributed cases.

| Disease | Sex | N | $N_{A_f}$ | Rank | $A_f$ | Rank $A_f$ |
| --- | --- | --- | --- | --- | --- | --- |
| S00.8 Superficial injury of other parts of head | F | 171 | 39 | 135 | 0.23 | 44 |
| S92 Fracture of foot, except ankle | M | 168 | 38 | 138 | 0.22 | 46 |
| E21 Hyperparathyroidism and other disorders of parathyroid gland | F | 296 | 66 | 106 | 0.22 | 48 |
| I21 Acute myocardial infarction | F | 1125 | 243 | 28 | 0.22 | 50 |
| M79.6 Pain in limb | F | 920 | 198 | 41 | 0.22 | 51 |
| M17 Gonarthrosis [arthrosis of knee] | F | 3623 | 735 | 3 | 0.20 | 57 |

## Discussion

### Effect of treatment on the treated (ETT)

An alternative attribution fraction, that is of more interest to clinicians or an individual, is the chance of having avoided a disease if you had not been exposed, but were subjected to the same confounding factors that you would have otherwise experienced. This situation is equivalent to estimating the “effect of treatment on the treated” (ETT) [1, 7], but the “treatment” is an exposure to smoking or BMI. For the situation considered here of figure 1, this counterfactual question can be formulated and expressed in terms of observational quantities in a similar way to before. The argument below considers the simpler situation of smokers versus never smoked, or max BMI tertile versus a lower BMI tertile, denoting exposed by *X* = *x*_1_ and unexposed by *X* = *x*_0_. Using counterfactual notation where 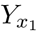 indicates the disease status of (e.g.) smokers, and 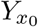 the disease status of non-smokers, then the ETT is defined as [1, 7],

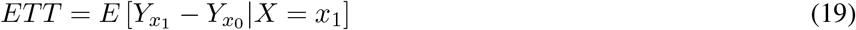

that can be thought of as estimating the difference between disease risk in smokers and non-smokers, when subjected to the same correlated confounding influences as smokers would experience. Following a previous derivation [7], and incorporating the same proportional hazards assumptions as before, this can be written as,

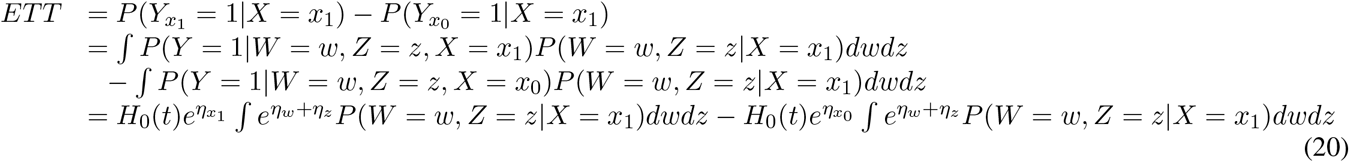

where the second term on the second line is usually justified with the backdoor adjustment formula Eq. 1, but corresponds to estimating the probability of disease when *X* = *x*_0_ but all other exposures are as they would have been if *X* = *x*_1_, and the third line uses the approximation of sufficiently rare diseases that the cumulative distribution function can be approximated by the cumulative hazard. Continuing to take the baseline value 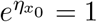, and dividing by the first term to get an attribution fraction, then gives,

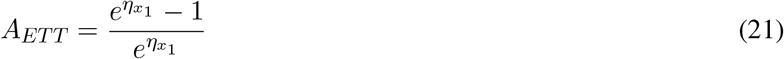

which solely involves the relative risk 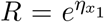 for e.g. smoking status, and is the simplest attribution fraction that occurs in the literature.

Because survival analyses are designed to estimate the influence of risk on an individual, with hindsight, perhaps Eq. 21 should not have been a surprise? Within the proportional hazards model, smoking will modify your risk of disease, independently of whether any other factors also do. From a population perspective, disease risk is determined by the overall combination of exposures, that will usually be correlated. This is why the attribution fraction for the population needs a more careful estimation that accounts for correlations between the exposures and confounding variables.

### Attribution fractions for causal estimates

Attribution formulae similar to those used here have existed in published literature since at least 1998 [5]. One aim of this paper is to emphasise that for a given causal model such as that in figure 1, the attribution fractions can only be used with a restricted range of potential risk modifiers, whose associations have a causal interpretation. If the causal model is incorrect, then the adjustment for confounding, and resulting estimates, are also likely to be incorrect. Alternately, if the measurement is too imprecise e.g. socio-economic status is likely to capture the influence of several factors that may include exposure to pollution, poor quality diet, poor living and working conditions, etc, then it may not be possible to estimate a meaningful causal association - for example, someone with an equivalent socio-economic status in a different country would experience different exposures and have different causal factors that influence their health. Another observation is that it may not be possible to obtain estimates of causal associations from a single analysis, but it might be possible to use the causal diagram to design an analysis that can estimate the parameters you are interested in. For example, changes in systolic blood pressure (SBP) can be caused by smoking or BMI, and therefore SBP should not be adjusted for if we are interested in the influence of smoking and BMI on disease risk. In contrast, if our interest was in SBP, then we would need to adjust for BMI and smoking if they can modify disease risk in any way other than through changes in SBP. These remarks have important consequences for meta-analyses of observational data, because different adjustments for covariates can equate to very different causal models, and could contribute a non-random source of heterogeneity.

### Attribution of disease to smoking and BMI

The attribution fractions for smoking, and BMI, are very heterogeneous, and can involve a reduction in risk (table 2). This highlights a difficulty in optimising lifestyle and drug treatments - changes in lifestyle or medication are likely to have a very heterogeneous influence on disease risk, with some risks lowered but others potentially increased. Another observation was that some of the associations were extremely strong, for example with *A*_*f*_ *>* 0.5. Strong germline genetic risk factors are often described as pathogenic when they substantially increase your risk of disease, and it seems reasonable to describe the influence of risk factors on a population as pathogenic when *A*_*f*_ is large, such as *A*_*f*_ *>* 0.5. For diseases estimated to have *A*_*f*_ *>* 0.5, eliminating the risk factors would be estimated to prevent the majority of those diseases in an equivalent population.

Attribution fractions can identify diseases for which lifestyle changes are likely to have the greatest impact. From a population perspective, eliminating smoking and controlling BMI in an equivalent population would be expected to avoid: the majority of diseases with *A*_*f*_ *>* 0.5 (red in table 1), between one third and one half of diseases with 0.35 *> A*_*f*_ *>* 0.5 (orange in table 1), between one fifth and one third of diseases with 0.2 *> A*_*f*_ *>* 0.35 (yellow in table 1). This slightly ad-hoc categorisation provides an indication of how the patterns of disease would be expected to change if smoking were eliminated and BMI were controlled in a population that was otherwise similar to that in UK Biobank.

Attributable fractions for a population will be larger if more of the population are exposed to a harmful risk factor. The Supplementary Material considers an example with a binary exposure X that is uncorrelated with W and confounders Z, and shows that provided *p*(*R −* 1) ≪ 1, where *R* is the relative risk and *p* is the proportion of the population that are exposed, then *A*_*f*_ ≃ *p*(*R −* 1). In that case, if the exposed proportion *p* were halved, then so would the attributable fraction. This highlights an important characteristic of Eqs. 8 and 13 - they measure the proportion of disease in a population that is attributed to an exposure. However, a clinician might be more interested in the proportion of disease in smokers is attributable to smoking, and an individual might be more interested in whether smoking substantially changes their risk of serious disease or death. Questions that refer to individuals can be tackled with counterfactual arguments and Eq. 21. An alternative approach is to consider the “probability of necessity” [1, 7], that is intended to assess whether it is more probable than not, that the disease would not have occurred if you had not been exposed to e.g. smoking. Such approaches allow specific individual cases to be assessed, but do not provide an overall characterisation of an exposure’s influence on population health.

When considering attribution fractions for smoking and BMI together, the study included diseases with statistically significant associations with *either* smoking or BMI. In this situation, especially when the number of cases are few, estimates for one of the two parameters can in principle be large and imprecise. This could produce misleading estimates for the joint attribution fraction of both smoking and BMI. An example is the strong protective association of smoking with Parkinson’s disease (table 4 in the Supplementary Material), that was substantially weakened by the association with BMI (table 2), even though the association with BMI was not statistically significant. This appears to be an isolated example, and the potential problem is less likely with more cases, but it highlights the importance of also considering the attribution fractions for each exposure separately.

### Limitations of the study

A limitation of this and related epidemiological studies, is that they consider the onset of new disease, unconfounded by prior disease. This is distinct from the more complex problem of determining the overall causal influence of smoking or BMI on the total incidence of diseases, that might involve sequences of disease incidence and repeated disease events. It also differs from the problem of determining how much disease is attributable to a combination of all exposures that are known to modify disease risk [18]. The advantages of this study over conventional epidemiological studies, is that it combines results from causal inference with observational data to allow the estimation of causal influences, and surveys a range of the most common diseases in UK Biobank. An unintended benefit of studying onset of new disease, unconfounded by prior disease, is that it tends to involve younger ages of incidence than average. For younger ages *t* with smaller *H*(*t*), the approximation *F* (*t*) ≃ *H*(*t*) will be better.

The causal diagram in figure 1 assumed that confounders such as socio-economic status influence both disease risk and the status of risk factors such as smoking status or BMI, but the main influence of risk factors such as smoking or BMI are directly on health. Although the causal diagram is plausible for the established risk factors and confounding factors considered here (see Methods for details), it would be prudent to check the causal and statistical assumptions in more detail before forming biological conclusions or modifying public health policy regarding a specific disease. To do this for all the diseases surveyed here would require a much more comprehensive study involving a wide range of clinicians with different specialities. That was beyond the scope of this study, whose primary aims are to develop a methodology and then use it to survey the likely influence of smoking and BMI on the overall pattern of disease incidence. Reassuringly however, the Supplementary Material’s figure 2 shows that the estimates were comparatively weakly modified by the choice of causal model. This suggests that if the model were incorrect for some of the diseases studied in this (UK Biobank) cohort, it is unlikely that the general patterns of attribution for different classes of diseases will substantially change. In fact, when the causal model is uncertain, it might still be possible to estimate attribution fractions if it can be shown that they are insensitive to the choice of causal model.

The proportional hazards modelling was intended to adjust for the strongest established risk factors and confounding factors, but it could also be improved by a more careful study for each disease. For example, this might require inclusion of time-dependent covariates, interactions, and e.g. “pack years smoking” in place of smoking status. The study here is also limited to UK Biobank data, that is an imperfect representation of the UK population.

The attribution fraction defined in Eq. 5 is dependent on age *t*, within which it assesses how much a risk factor has modified the population’s risk of disease. In practice this can often be approximated by Eq. 10, but an alternative is to use the more theoretical definition of attribution fraction for a sufficiently low age *t* → 0 that Eq. 5 exactly equals Eq. 10. This latter definition provides a measure of attribution that is independent of age, and independent of the specific epidemiological study (e.g. a study might consider only the first disease observed by an individual, and unconfounded by prior disease). It has the added practical benefit that it can be estimated using well-understood proportional hazard models, and in practice for most of the average human lifespan the differences between Eq. 5 and 10 are expected to be small for most diseases in most people. Examples in the Supplementary Material suggest that for relative risks greater than 1, even for examples with the most extreme deviations, the approximation of Eq. 10 is often reasonable until 80 or 90 years, with the attribution fraction slowly decreasing with age. For all of these reasons, the age-independent Eq. 10 that was used here may often be the most appropriate definition.

### Conclusions

The aim was to characterise and classify the causal influence of established risk factors on common diseases, using observational data from UK Biobank. Assuming a simple causal model (figure 1), the theory of causal inference can allow the estimation of causal associations from observational data. This is possible for some, but not all of the risk factors that are usually included in epidemiological studies, but includes smoking and BMI. The “backdoor criteria” from causal inference was used to derive a population attribution fraction, that can be approximated with a proportional hazards model. The proportional hazards approximation Eq. 10 can be interpreted as the attribution fraction for disease occurring by age *t*, in the theoretical limit of *t* → 0 (a sufficiently low age for the approximation to be accurate). This defines an attribution fraction that is easy to evaluate with well-understood epidemiological methods, and is independent of age. In practice, for most of a typical UK human lifespan, the age-dependent differences between Eq. 5 and Eq. 10 are expected to be small for most diseases in most people. Conventional epidemiological methods were used to estimate associations between established risk factors and common diseases in UK Biobank data. These (proportional hazard) estimates were used to evaluate causal attribution fractions for smoking and BMI on the incidence of 226 diseases, identifying the diseases and ICD-10 chapter disease classifications whose risks were the most modifiable. The results suggest which diseases and classes of diseases in the UK Biobank cohort are likely to be most strongly influenced by smoking and BMI, and provides a template for more comprehensive future studies. Although the studies here involved established risk and confounding factors, biologically-informed clinical input is needed to check the causal and statistical assumptions before forming any biological conclusions, or modifying public health policy about a specific disease.

## Data Availability

The study uses UK Biobank data, that is available by application from: www.ukbiobank.ac.uk

## Data availability

UK Biobank data can be accessed by application through www.ukbiobank.ac.uk, and summary data produced during this study is available at the link provided below: https://osf.io/dsgxw/

UK Biobank has approval by the Research Ethics Committee (REC) under approval number 16/NW/0274. UK Biobank obtained participant’s consent for the data to be used for health-related research, and all methods were performed in accordance with the relevant guidelines and regulations.

## Code availability

R code used to produce figures from summary data is available from the link below: https://osf.io/dsgxw/

The full code for use with non-summary data will be returned with other results to UK Biobank (see www.ukbiobank.ac.uk).

## Acknowledgements

Thank you to Professor Robert Clarke for suggesting a sensitivity analysis to strengthen the results, and to Professor Peter Sasieni for critical and constructive comments on an earlier draft. This research has been conducted using data from UK Biobank, a major biomedical database, under application number 42583. Anthony Webster was supported by an intermediate research fellowship from the Nuffield Department of Population Health (NDPH), University of Oxford.

