## Supplemental Material for "Causal attribution fractions, and the attribution of smoking and BMI to the landscape of disease incidence in UK Biobank"

### A Attribution fractions

#### A.1 Bounds on attribution fractions

By definition, if the relative risks $e^{\eta_x}$ and $e^{\eta_w+\eta_z}$ are positively correlated, then $E[e^{\eta_x}e^{\eta_w+\eta_z}] - E[e^{\eta_x}]E[e^{\eta_w+\eta_z}] \geq 0$, where $E$ is used to denote expectations. Therefore, using $\exp(s) \geq 0$ for any real-valued $s$, we can rearrange the inequality as,

$$\begin{array}{ll} E[e^{\eta_x}e^{\eta_w+\eta_z}] & \geq E[e^{\eta_x}]E[e^{\eta_w+\eta_z}] \\ \frac{1}{E[e^{\eta_x}]} & \geq \frac{E[e^{\eta_w+\eta_z}]}{E[e^{\eta_x}e^{\eta_w+\eta_z}]} \\ 1 - \frac{E[e^{\eta_w+\eta_z}]}{E[e^{\eta_x}e^{\eta_w+\eta_z}]} & \geq 1 - \frac{1}{E[e^{\eta_x}]} \end{array} \tag{1}$$

Eq. 1 shows that if the relative risks for $x$, $w$, and $z$ are positively correlated, then the attributable fraction for disease risk within the population, is greater than would be estimated using the average relative risk, with estimates using the mean relative risk providing a lower bound. If $x$ and $z$ are negatively correlated then the $\geq$ sign is be replaced by $\leq$.

Another quantity that might be considered is the expected value of the attributed fraction $1 - 1/e^{\eta_x}$, that is $E[1 - 1/e^{\eta_x}] = 1 - E[1/e^{\eta_x}]$. Because $1/e^{\eta_x}$ is concave, Jensen's inequality gives,

$$E\left[\frac{1}{e^{\eta_x}}\right] \geq \frac{1}{E[e^{\eta_x}]} \tag{2}$$

and as a result,

$$1 - \frac{1}{E[e^{\eta_x}]} \geq 1 - E\left[\frac{1}{e^{\eta_x}}\right] = E\left[1 - \frac{1}{e^{\eta_x}}\right] \tag{3}$$

If $e^{\eta_x}$ and $e^{\eta_w+\eta_z}$ are positively correlated, then Eqs. 3 and 1 indicate that $E[1 - 1/e^{\eta_x}]$ will also bound Eq. 10 of the main text. However, this would not be true if $e^{\eta_x}$ and $e^{\eta_w+\eta_z}$ were negatively correlated.

#### A.2 Relation to other attributable fractions

A recent study[1] with a proportion $p$ exposed to a virus, and an estimated relative risk $R$, reported an attribution fraction of $A = p(R-1)/R$. Here it is briefly outlined when this will approximate Eq. 10 of the main text. Assume that $e^{\eta_x}$, $e^{\eta_w}$, and $e^{\eta_z}$ are uncorrelated, so that with the approximation $F(t) \simeq H(t)$, Eq. 8 in the main text simplifies to Eq. 14 in the main text, that may be approximated as,

$$A_f \simeq 1 - \frac{1}{\frac{1}{n}\sum_{i=1}^{n} e^{\eta_{x_i}}} \tag{4}$$

If we consider a proportion $p$ that are exposed with relative risk $R$, and a proportion $(1-p)$ that are unexposed, then using Eq. 4,

$$\begin{array}{ll} A_f & \simeq 1 - \frac{1}{(1-p)+pR} \\ & = \frac{p(R-1)}{1+p(R-1)} \\ & = p\left(1 - \frac{1}{R}\right)\left(\frac{R}{1+p(R-1)}\right) \\ & \simeq p\left(1 - \frac{1}{R}\right) \end{array} \tag{5}$$

where the approximation in the final line follows if $R-1$ is small enough, as it often can be. A better approximation follows from the second line, where $p(R-1) \ll 1$ ensures that $A_f \simeq p(R-1)$. If $p \simeq 1$, then $A_f \simeq (R-1)/R$ as usual, as can be seen from the first or second line above.

### B Unmeasured confounders and mediation - the "frontdoor criteria"

Another important result from causal inference, is the "frontdoor criteria"[2,3]. A well-known example[3] is assessing the influence of smoking on disease risk in the presence of *unmeasured* confounders that influence both smoking use and disease risk, by using an additional measurement of tar in peoples' lungs (figure 1). Again we consider the adjustment formula for this situation in the limit of rare diseases, as above, and consider the simple specific example with continuous variables for e.g. average number of cigarettes per day and tar content of lungs. Although the estimated incidence rates will differ from those using proportional hazards models, the causal estimate for the influence of smoking on lung cancer, is the same as we might (with hindsight) have anticipated from mediation studies.

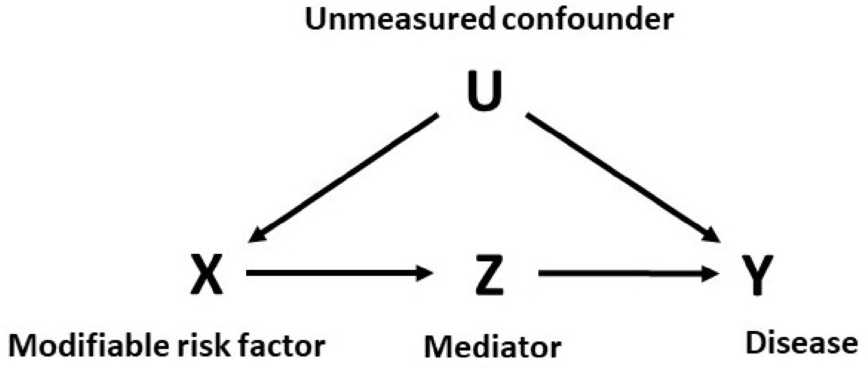


**Figure 1.** The "frontdoor criteria" estimates the causal influence of an exposure $\text{do}(X=x)$, that is mediated by $Z$, in the presence of unmeasured confounders $U$ that influence both the disease risk and the exposure $X$.

For the situation described in figure 1, the "front door" adjustment formula states[2,3],

$$P(Y=y|\text{do}(X=x)) = \sum_z \sum_{x'} P(Y=y|Z=z, X=x')P(X=x')P(Z=z|X=x) \tag{6}$$

Using this, and proceeding as before,

$$\begin{aligned} P(Y=1, T<t|\text{do}(X=x)) &= \sum_z \sum_{x'} P(Y=1, T<t|X=x', Z=z)P(Z=z|X=x)P(X=x') \\ &\simeq \sum_z \sum_{x'} e^{\eta_{x'}} e^{\eta_z} H_0(t) P(Z=z|X=x)P(X=x') \\ &= H_0(t)\left(\sum_z e^{\eta_z} P(Z=z|X=x)\right)\left(\sum_{x'} e^{\eta_{x'}} P(X=x')\right) \end{aligned} \tag{7}$$

Next consider the specific example where $\eta_{x'} = \beta_x x'$, $\eta_z = \beta_z z$, $P(X=x)$ is a normal distribution $N(\mu_x, \sigma_x^2)$, and $P(Z=z|X=x)$ is a normal distribution $N(\alpha x, \sigma_z^2)$, where in the latter case $\alpha$ is a constant and the mean of $z$ is $\alpha x$. Understanding that the sums should be considered as integrals when variables are continuous, then we have,

$$\sum_{x'} e^{\eta_{x'}} P(X=x') = \exp(\beta_x \mu_x) \exp\left(\frac{\sigma_x^2 \beta_x^2}{2}\right) \tag{8}$$

and,

$$\sum_z e^{\eta_z} P(Z=z|X=x) = \exp(\beta_z \alpha x) \exp\left(\frac{\sigma_z^2 \beta_z^2}{2}\right) \tag{9}$$

giving,

$$P(Y=1, T<t|\text{do}(X=x)) \simeq H_0(t) \exp(\beta_x \mu_x) \exp\left(\frac{\sigma_x^2 \beta_x^2}{2} + \frac{\sigma_z^2 \beta_z^2}{2}\right) \exp(\beta_z \alpha x) \tag{10}$$

The incidence rate at baseline $X = x_0$ is determined by the first three terms, and differs from a proportional hazard estimate that is adjusted by either or both, of $x$ or $z$. The first two terms are equivalent to a proportional hazards estimate with $x$ at the mean exposure $\mu_x$ and $z$ at the baseline value, and the third term quantitatively accounts for the spread in values of $x$ and $z$ about their mean values. The influence of $\text{do}(X=x)$, is seen in the last term $e^{\beta_z \alpha x}$, with the change in risk being mediated by $z$ in a very simple and intuitive way.

For the situation considered here, where there is solely an indirect effect of the exposure through the mediator, this estimate is the same as for a mediation analysis with *measured* confounding[4]. Interestingly, in the equivalent mediation analysis with measured confounding, the influence of measured confounding on the estimate[1], does not appear in the resulting expressions for natural direct, and indirect, effects. This appears to explain the agreement between estimates with measured, and unmeasured confounding - for the model of figure 1 in limit of rare diseases and a proportional hazards model, the estimate is (apparently) unaffected by confounding.

Equation 7 applies to any situation described by figure 1, and the example given can be generalised, e.g. to multivariate normal distributions.

[1]For a solely indirect effect, $\gamma_1 = \gamma_3 = 0$ in Eq. 4.6 on page 101 of[4], and measured confounding is accounted for through the coefficient $\gamma_4$, that does not subsequently appear in the equations for natural direct and indirect effects.

### C Relative risks

Using the same approximations used to derive Eq. 4 of the main text, it can be written in several ways, for example,

$$\begin{aligned}&P(Y=1,T<t|\mathrm{do}(X=x))\\&=e^{\eta_x}A_Z H_0(t)\\&=e^{\eta_x}A_Z P(Y=1,T<t|X=x_0,Z=z_0)\\&=A_Z P(Y=1,T<t|X,Z=z_0)\end{aligned} \tag{11}$$

When education and socio-economic factors are represented by $Z$, then the factor $A_Z$ accounts for changes in risk due to both socio-economic factors and education, and the influence of setting $X=x$ is calculated through the factor $e^{\eta_x}$. If we could set $X$ equal to the baseline values $x_0$, the probability distribution would be proportional to the baseline hazard function $H_0(t)$, amplified or shrunk by the factor $A_Z$. If the baseline values corresponded to the lowest disease risk, then $A_Z H_0(t)$ would be the lowest possible disease incidence rate that could have been achieved through lifestyle changes. Eq. 11 can be written as,

$$\frac{P(Y=1,T<t|\mathrm{do}(X=x))}{P(Y=1,T<t|\mathrm{do}(X=x_0))}=e^{\eta_x} \tag{12}$$

This shows that the relative risk of disease within time $t$ for a population with $X=x$, compared with a population with baseline values of $X=x_0$, is equal to the relative risk from observational studies, that have,

$$\frac{h(t|X=x,Z=z_0)}{h(t|X=x_0,Z=z_0)}=e^{\eta_x} \tag{13}$$

### D Supplementary tables and figures

#### D.1 Comparison between attribution fractions

Figure 2 compares attribution fractions estimated using Eqs. 10 and 14 of the main text (Eq. 14 is the equivalent estimate to that used by the World Health Organisation[8]).

#### D.2 Estimates solely involving smoking or BMI

Figures 1-4, show equivalent tables and plots to those in the main text, but for attribution fractions solely due to BMI or smoking alone.

#### D.3 The estimate $F(t) \simeq H(t)$

Probability densities $F(t)$ for 400 diseases in men and women, without confounding by prior disease, were modelled with Weibull distributions[5]. For age groups of 60, 70, 80, and 90 years, figure 4 provides histograms for the number of diseases having occurred with a given probability interval, and the cumulative proportion of diseases included by that interval. Even at age 90, almost all diseases had an estimated probability of less than 0.2, for which values the estimate $F(t) \simeq H(t)$ is very good. With confounding by prior disease, or with greater than average risk factors, the probabilities would be higher.

Unfortunately the approximation $F(t) \simeq H(t)$ becomes less reliable when individuals have risk factors that lead to a much higher relative risk than the general population. To explore where the approximations start to fail, examples with relative risks of 1.1, 2.0, and 5.0 are considered, for a late-onset disease whose risk increases rapidly in later life and for a sporadic disease whose risk is moderate throughout life but increases comparatively slowly with age[5]. The diseases were modelled with a Weibull distribution with survival function $S(t) = \exp(-e^{\eta}(t/L)^k)$, where $\eta$ is a linear predictor for adjustment so $e^{\eta} \equiv RR$ is the relative risk, $L$ is a parameter that sets a scale for age $t$, and $k$ is a dimensionless parameter. For the late-onset disease example we took $L = 115$ and $k = 6.8$, and for the sporadic disease example we took $L = 190$ and $k = 2.0$. The population will contain a mixture of individuals with relative risks ranging in values, some of which may be less than one.

Figure 5 compares $F(t)$ with its approximation by $F(t) \simeq H(t)$, and also explores how the approximation modifies the estimated attribution fractions (if defined by Eq. 8 instead of Eq. 10, both from the main text). Note that many individuals in a population will often have small relative risks, and the overall combination of relative risks from within the population will

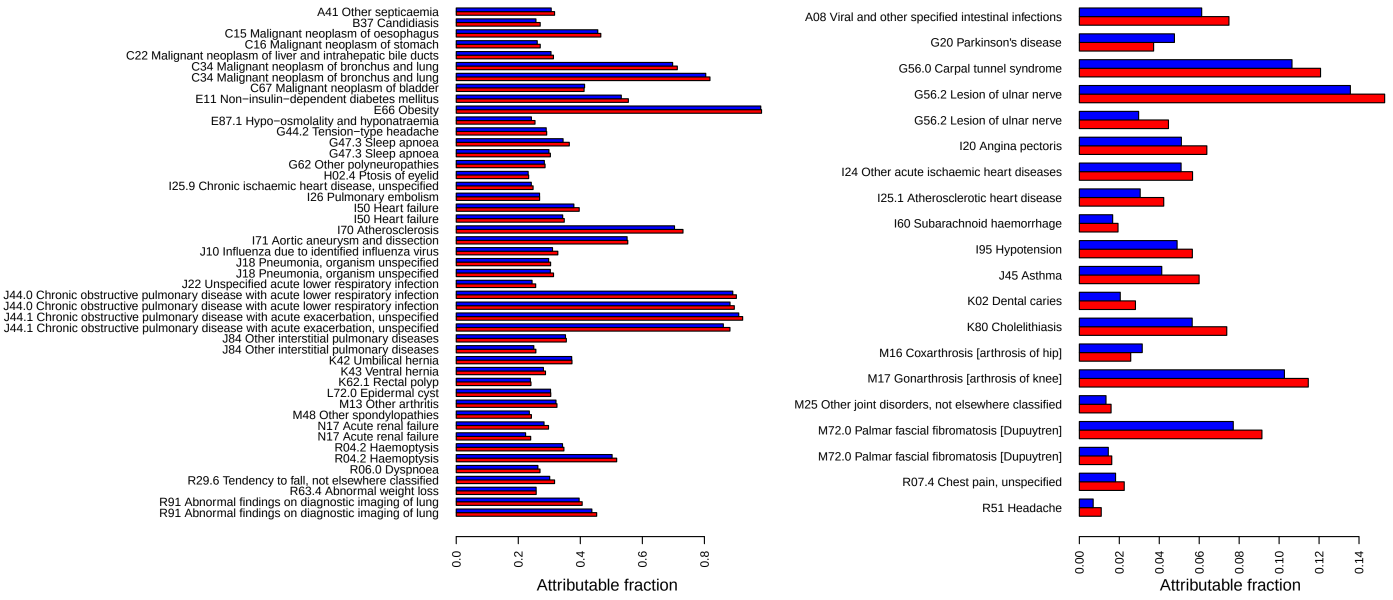


**Figure 2.** Attributable fractions ($A_f$) for smoking and BMI, estimated with Eqs. 10 (red) and 14 (blue) of the main text. Eq. 14 is equivalent to the World Health Organisation's[8] $A_f$. Eq. 10 from the main text estimates causal associations, that accounts for correlations between risk factors and confounders. Left plot: diseases with the highest 25% $A_f$. Right plot: diseases where Eqs. 10 and 14 differed the most.

| Disease | Sex | N | $N_{A_f}$ | Rank | $A_f$ | Rank $A_f$ |
|---|---|---|---|---|---|---|
| E66 Obesity | F | 311 | 305 | 7 | 0.98 | 1 |
| E11 Non-insulin-dependent diabetes mellitus | M | 206 | 94 | 38 | 0.45 | 2 |
| K42 Umbilical hernia | F | 297 | 120 | 32 | 0.40 | 3 |
| G47.3 Sleep apnoea | F | 381 | 121 | 31 | 0.32 | 4 |
| I50 Heart failure | M | 359 | 111 | 34 | 0.31 | 5 |
| G62 Other polyneuropathies | M | 175 | 47 | 66 | 0.27 | 6 |
| M13 Other arthritis | M | 215 | 57 | 53 | 0.27 | 7 |
| I50 Heart failure | F | 280 | 74 | 44 | 0.26 | 8 |
| I26 Pulmonary embolism | F | 836 | 194 | 16 | 0.23 | 9 |
| G47.3 Sleep apnoea | M | 776 | 172 | 21 | 0.22 | 10 |
| J22 Unspecified acute lower respiratory infection | F | 1506 | 326 | 6 | 0.22 | 11 |
| J10 Influenza due to identified influenza virus | F | 211 | 43 | 73 | 0.20 | 12 |
| I25.9 Chronic ischaemic heart disease, unspecified | M | 254 | 51 | 59 | 0.20 | 13 |
| M81 Osteoporosis without pathological fracture | F | 946 | 191 | 17 | 0.20 | 14 |

**Table 1.** BMI only: Sex indicates diseases in males (M) or females (F), $N$ are total cases, $N_{A_f}$ are cases attributed to BMI, $A_f$ is the attributable fraction for deviations from the mid-tertile of BMI. For obesity, unsurprisingly $A_f \simeq 1$. Reporting errors may have prevented $A_f = 1$ for obesity.

| Disease | Sex | N | $N_{A_f}$ | Rank | $A_f$ | Rank $A_f$ |
|---|---|---|---|---|---|---|
| J44.1 Chronic obstructive pulmonary disease with acute exacerbation, unspecified | F | 209 | 189 | 28 | 0.90 | 1 |
| J44.0 Chronic obstructive pulmonary disease with acute lower respiratory infection | F | 416 | 368 | 10 | 0.89 | 2 |
| J44.0 Chronic obstructive pulmonary disease with acute lower respiratory infection | M | 417 | 364 | 12 | 0.87 | 3 |
| J44.1 Chronic obstructive pulmonary disease with acute exacerbation, unspecified | M | 261 | 227 | 21 | 0.87 | 4 |
| C34 Malignant neoplasm of bronchus and lung | M | 1018 | 811 | 2 | 0.80 | 5 |
| I70 Atherosclerosis | M | 156 | 115 | 50 | 0.74 | 6 |
| C34 Malignant neoplasm of bronchus and lung | F | 996 | 702 | 3 | 0.70 | 7 |
| I71 Aortic aneurysm and dissection | M | 402 | 230 | 20 | 0.57 | 8 |
| R04.2 Haemoptysis | M | 314 | 134 | 43 | 0.43 | 9 |
| C15 Malignant neoplasm of oesophagus | M | 473 | 194 | 26 | 0.41 | 10 |
| R91 Abnormal findings on diagnostic imaging of lung | M | 358 | 145 | 37 | 0.41 | 11 |
| C67 Malignant neoplasm of bladder | M | 1063 | 414 | 8 | 0.39 | 12 |
| R91 Abnormal findings on diagnostic imaging of lung | F | 372 | 131 | 45 | 0.35 | 13 |
| J84 Other interstitial pulmonary diseases | F | 177 | 53 | 83 | 0.30 | 14 |
| B37 Candidiasis | M | 173 | 48 | 90 | 0.28 | 15 |
| J84 Other interstitial pulmonary diseases | M | 234 | 64 | 72 | 0.27 | 16 |
| J90 Pleural effusion, not elsewhere classified | M | 524 | 137 | 41 | 0.26 | 17 |
| R04.2 Haemoptysis | F | 239 | 62 | 75 | 0.26 | 18 |
| G56.2 Lesion of ulnar nerve | F | 222 | 52 | 85 | 0.23 | 19 |
| K92.0 Haematemesis | M | 156 | 36 | 102 | 0.23 | 20 |
| C22 Malignant neoplasm of liver and intrahepatic bile ducts | M | 171 | 39 | 97 | 0.23 | 21 |
| R29.6 Tendency to fall, not elsewhere classified | M | 182 | 40 | 94 | 0.22 | 22 |
| R06.0 Dyspnoea | M | 650 | 142 | 39 | 0.22 | 23 |
| J18 Pneumonia, organism unspecified | M | 3011 | 645 | 4 | 0.21 | 24 |
| C16 Malignant neoplasm of stomach | M | 260 | 55 | 79 | 0.21 | 25 |
| H02.0 Entropion and trichiasis of eyelid | M | 337 | 68 | 69 | 0.20 | 26 |
| K62.1 Rectal polyp | M | 1143 | 231 | 18 | 0.20 | 27 |
| G44.2 Tension-type headache | F | 152 | 31 | 110 | 0.20 | 28 |

**Table 2.** Smoking only: Sex indicates diseases in males (M) or females (F), $N$ are total cases, $N_{A_f}$ are cases attributed to smoking, $A_f$ is the attributable fraction.

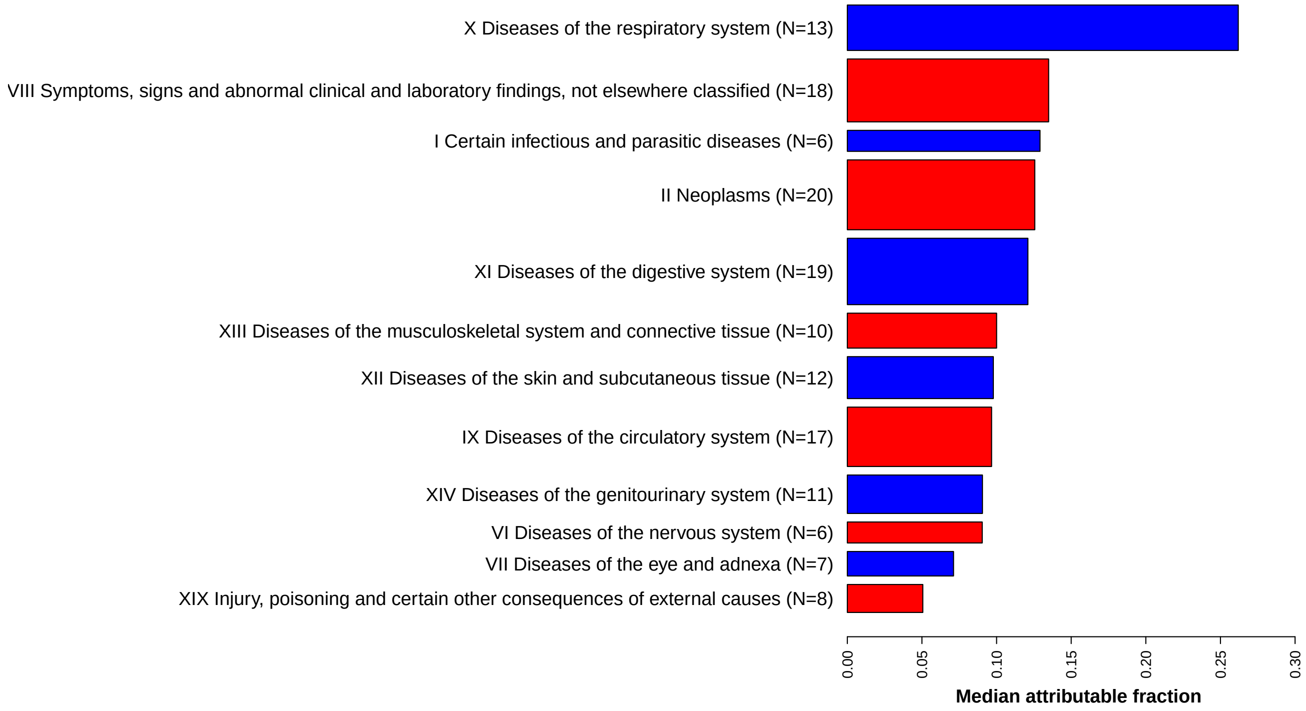


**Attribution fractions for smoking (existing UK Biobank population, versus if all never smoked)**

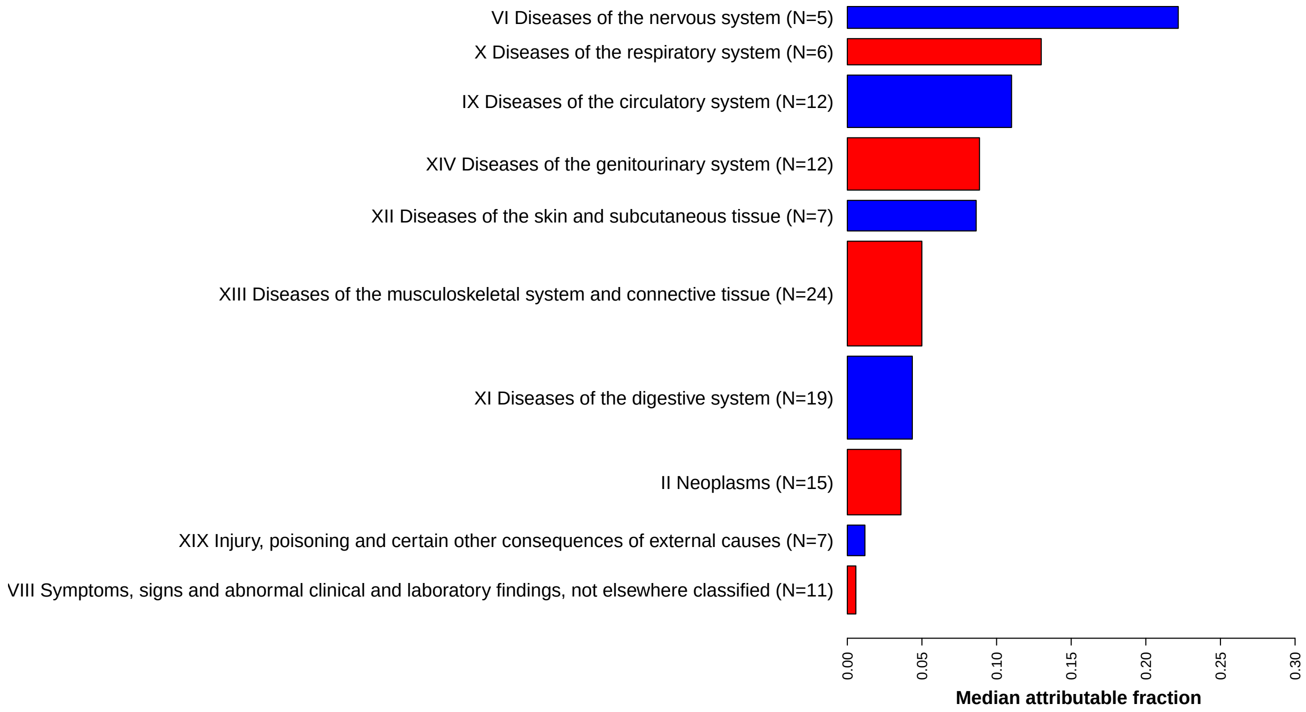


**Attribution fractions for BMI (existing UK Biobank population, versus if all were mid-tertile BMI)**

**Figure 3.** Median attributable fractions for each ICD-10 chapter with at least 5 diseases where $A_f > 0.2$. Bar widths are proportional to the number of diseases in each chapter.

| Disease | Sex | N | $N_{A_f}$ | Rank | $A_f$ | Rank $A_f$ |
|---|---|---|---|---|---|---|
| N81 Female genital prolapse | F | 4199 | −788 | 1 | −0.190 | 1 |
| L72.0 Epidermal cyst | F | 525 | −96 | 13 | −0.180 | 2 |
| N41 Inflammatory diseases of prostate | M | 572 | −103 | 12 | −0.180 | 3 |
| S02 Fracture of skull and facial bones | M | 355 | −36 | 23 | −0.100 | 4 |
| K40 Inguinal hernia | F | 493 | −49 | 20 | −0.100 | 5 |
| R19.8 Other specified symptoms and signs involving the digestive system and abdomen | F | 302 | −30 | 25 | −0.098 | 6 |
| M20.1 Hallux valgus (acquired) | F | 2875 | −280 | 2 | −0.097 | 7 |
| S76.1 Injury of quadriceps muscle and tendon | M | 215 | −20 | 26 | −0.095 | 8 |
| R79 Other abnormal findings of blood chemistry | M | 1905 | −169 | 5 | −0.089 | 9 |
| D04 Carcinoma in situ of skin | M | 201 | −16 | 27 | −0.081 | 10 |
| K52.9 Non-infective gastro-enteritis and colitis, unspecified | M | 1061 | −76 | 15 | −0.071 | 11 |
| S82 Fracture of lower leg, including ankle | F | 2163 | −110 | 10 | −0.051 | 12 |

**Table 3.** BMI only: Diseases with the strongest protective associations, ranked by attributable fraction ($A_f$). Sex indicates diseases in males (M) or females (F), $N$ are total cases, $N_{A_f}$ are the number of cases attributed to BMI.

| Disease | Sex | N | $N_{A_f}$ | Rank | $A_f$ | Rank $A_f$ |
|---|---|---|---|---|---|---|
| G20 Parkinson's disease | F | 152 | −31 | 18 | −0.200 | 1 |
| D03 Melanoma in situ | M | 272 | −44 | 14 | −0.160 | 2 |
| G20 Parkinson's disease | M | 289 | −41 | 15 | −0.140 | 3 |
| C43 Malignant melanoma of skin | M | 723 | −86 | 8 | −0.120 | 4 |
| S52 Fracture of forearm | M | 687 | −74 | 9 | −0.110 | 5 |
| K31.7 Polyp of stomach and duodenum | F | 870 | −89 | 7 | −0.100 | 6 |
| C54 Malignant neoplasm of corpus uteri | F | 878 | −70 | 12 | −0.080 | 7 |
| R79 Other abnormal findings of blood chemistry | M | 1905 | −150 | 4 | −0.079 | 8 |
| R19.5 Other fecal abnormalities | F | 544 | −32 | 17 | −0.059 | 9 |
| N40 Hyperplasia of prostate | M | 3928 | −214 | 3 | −0.055 | 10 |
| C61 Malignant neoplasm of prostate | M | 5800 | −304 | 1 | −0.052 | 11 |
| K31.7 Polyp of stomach and duodenum | M | 300 | −15 | 21 | −0.051 | 12 |

**Table 4.** Smoking only: Diseases with the strongest protective associations, ranked by attributable fraction ($A_f$). Sex indicates diseases in males (M) or females (F), $N$ are total cases, $N_{A_f}$ are the number of cases attributed to smoking.

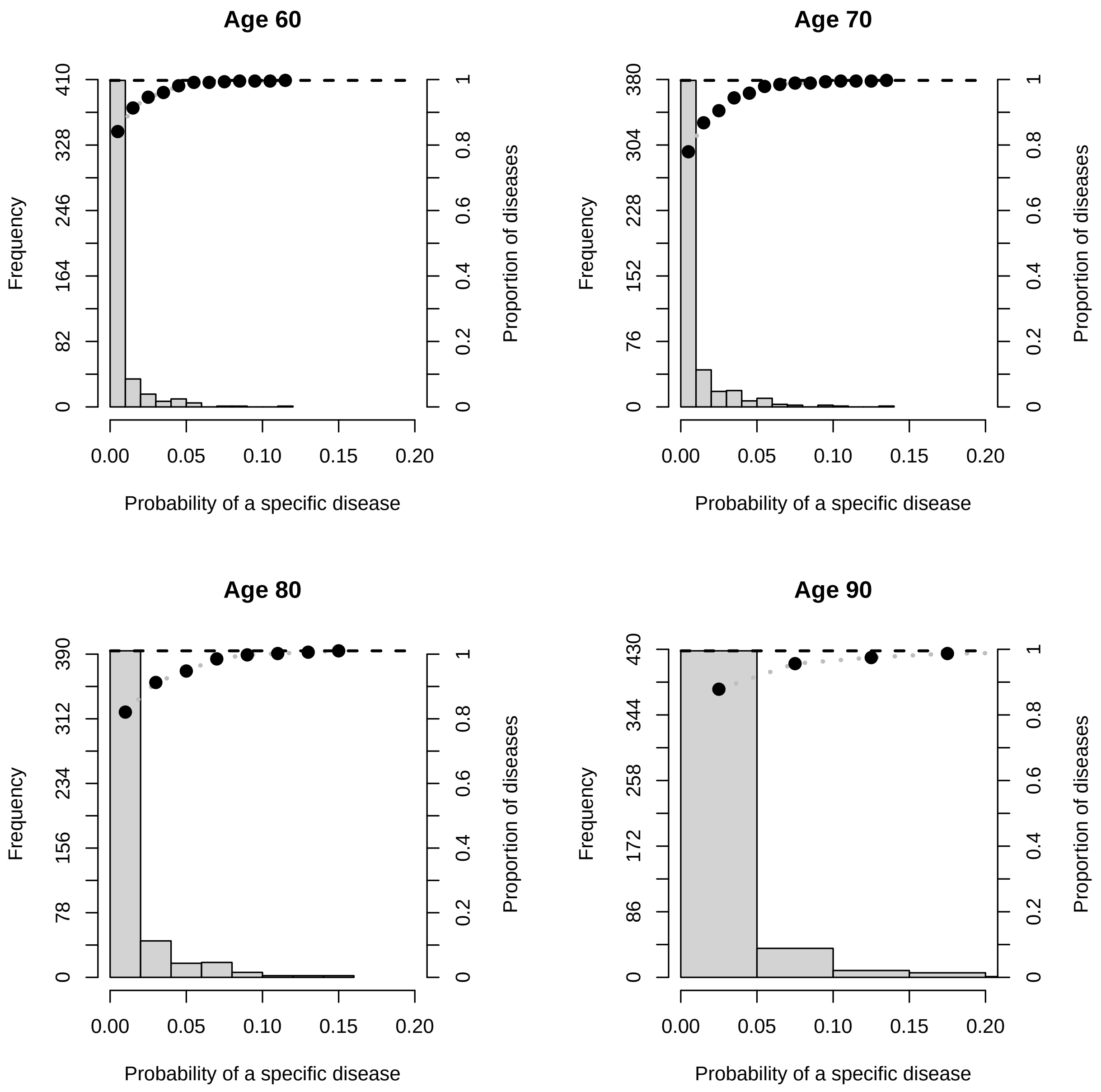


**Figure 4.** The probability densities $F(t)$ for 400 diseases in men and women were modelled with Weibull distributions[5]. For each age group of 60, 70, 80, and 90 years, the histogram shows the number of diseases with a given probability of having occurred (for no confounding by prior disease), and the right axes show the cumulative proportion of diseases.

determine the accuracy of Eq. 8's approximation of Eq. 10. For late-onset diseases, whose risk is small until late in life, the approximations are all very good until age 80, but start to fail for the larger relative risks from age 90 onwards. For the sporadic diseases and relative risks of 1.1 and 2.0, the approximations are fairly good until 80 or 90, but when the relative risk is 5.0 the approximation starts to fail from approximately age 60 onwards. Overall, for most individuals and diseases, the approximation $F(t) \simeq H(t)$ is adequate for average UK life expectancies of approximately 80 years, in other words, for most of a typical UK human lifespan. The strongest failure of the estimates are for individuals with large relative risks, and for diseases that are more likely to be observed earlier in life (sporadic diseases). For unadjusted fits and small relative risks, the estimate of $F(t) \simeq H(t)$ is good even for ages approaching 100. An alternative option suggested in the main text, is to regard Eq. 8 as an (age-independent) definition for ages $t \to 0$, which will be a reasonable approximation for most individuals, diseases, and ages up to the average UK life expectancy of approximately 80 years.

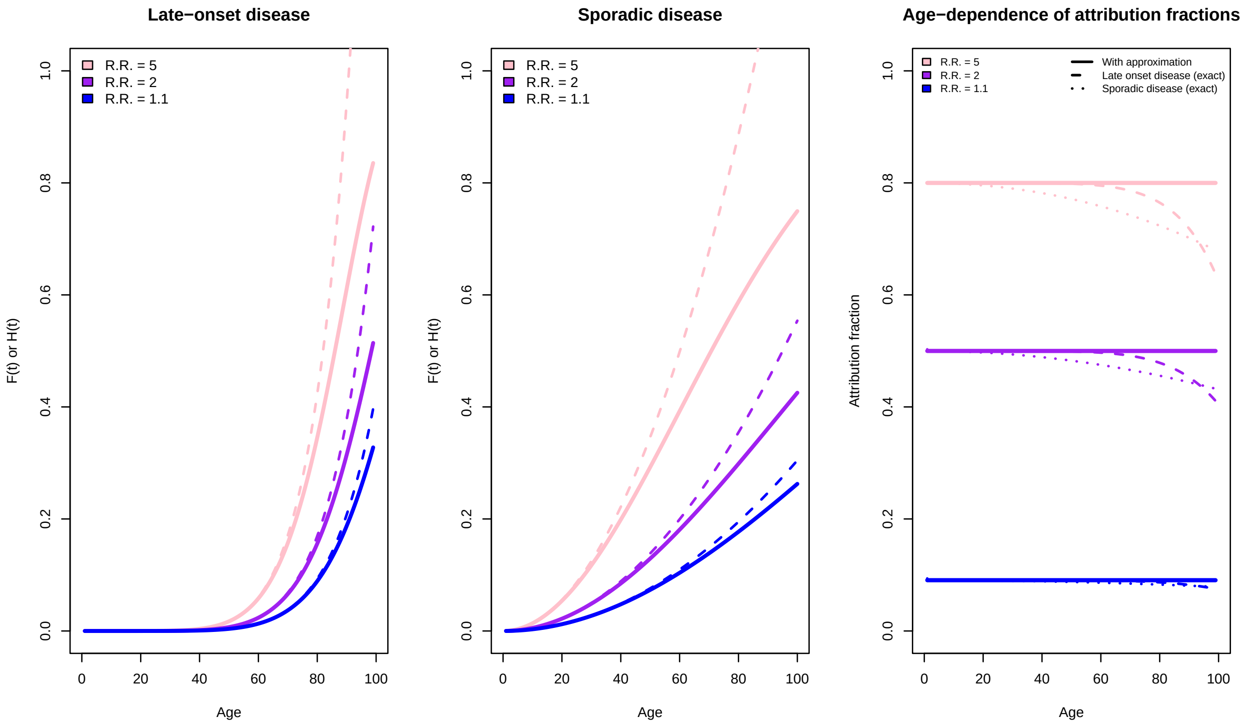


**Figure 5.** The left and central figures show how the approximation $F(t) \simeq H(t)$ starts to fail at large enough ages, and how larger relative risks (R.R.) make the approximation worse. Comparing the left and central figure, the approximation is much better for late-onset diseases that only occur in old age. The right figure shows how the approximation causes the age-dependent attribution fraction (Eq. 10 of the main text), to deviate from its age-independent approximation (Eq. 8 of the main text), at large enough ages. Even the largest deviations are within about 20% of the age-independent approximation (right figure).
